# Canadian Adaptive Platform Trial of Treatments for COVID in Community Settings (CanTreatCOVID): protocol for a randomized controlled adaptive platform trial of treatments for acute SARS-CoV-2 infection in community settings

**DOI:** 10.1101/2024.11.25.24317904

**Authors:** Benita Hosseini, Amanda Condon, Bruno R. da Costa, Peter Daley, Michelle Greiver, Peter Jüni, Todd C. Lee, Kerry McBrien, Emily G. McDonald, Srinivas Murthy, Peter Selby, Melissa Andrew, Kris Aubrey-Bassler, David Barber, Brendan Barrett, Chris Butler, Noah Crampton, Simone Dahrouge, Ali Damji, Robert Fowler, Stephanie Garies, Catherine Hudon, Jennifer Hulme, Jennifer Isenor, David Jenkins, Rosemarie Lall, Annie LeBlanc, Christine Leong, Paul Little, Aisha Lofters, Sarvesh Logsetty, Sylvain Lother, Marie-Thérèse Lussier, Laura MacLaren, Derelie Mangin, Emily Marshall, John Marshall, Rita McCracken, Rahim Moineddin, Briana Orava, Jean-Sebastien Paquette, Jay Jae Hee Park, Navindra Persaud, Valeria Rac, Vivian Ramsden, Jennifer Rayner, Diana Sanchez Ramirez, Lynora Saxinger, Haolun Shi, Alexander Singer, Rae Spiwak, Anita Srivastava, Abhimanyu Sud, Jean-Éric Tarride, Deanna Telner, Ross Upshur, Sakina Walji, Rachel Walsh, Machelle Wilchesky, Sabrina Wong, Brianne Wood, Ryan Zarychanski, Barbara Zelek, Yoav Keynan, Jolanta Pisczek, Daniel Warshafsky, Andrew D. Pinto

**Author notes:** Address correspondence to: Andrew D. Pinto Upstream Lab, MAP Centre for Urban Health Solutions, Li Ka Shing Knowledge Institute, Unity Health Toronto 30 Bond Street, Toronto, ON, M5B 1W8.

## Abstract

**Introduction:** While effective vaccines and natural immunity have significantly reduced hospitalizations and the need for critical care, SARS-CoV-2 is now endemic and is expected to continue to pose a threat to health. New variants are expected to continue to emerge, and vaccines may become less effective. Effective and affordable therapeutics for SARS-CoV-2 that can be easily used in community settings are needed to accelerate recovery, reduce hospitalizations and mortality, and mitigate the development of post-acute sequelae of SARS-CoV-2, also known as “long COVID.” In this paper we present the design of the Canadian Adaptive Platform Trial of Treatments for COVID in Community Settings (CanTreatCOVID).

**Methods and analysis:** CanTreatCOVID is an open-label, individually randomized, multi-centre, national adaptive platform trial designed to evaluate the clinical and cost-effectiveness of therapeutics for non-hospitalized SARS-CoV-2 patients across Canada. Eligible participants must present with symptomatic SARS-CoV-2 infection, confirmed by PCR or rapid antigen testing (RAT), within 5 days of symptom onset. The trial targets two groups that are expected to be at higher risk of more severe disease: (1) individuals aged 50 years and older, and (2) those aged 18-49 years with one or more comorbidities. CanTreatCOVID uses numerous approaches to recruit participants to the study, including a multi-faceted public communication strategy and outreach through primary care, out-patient clinics, and emergency departments. Participants are randomized to receive either usual care, including supportive and symptom-based management, or an investigational therapeutic selected by the Canadian COVID-19 Outpatient Therapeutics Committee. The first therapeutic arm evaluates nirmatrelvir/ritonavir (Paxlovid™), administered twice daily for 5 days. The second therapeutic arm investigates a combination antioxidant therapy (selenium 300 µg, zinc 40 mg, lycopene 45 mg, and vitamin C 1.5 g), administered for 10 days. The primary outcome is all-cause hospitalization or death within 28 days of randomization.

**Ethics and dissemination:** The CanTreatCOVID master protocol and sub-protocols have been approved by Health Canada and local research ethics boards in the participating provinces across Canada. The results of the study will be disseminated to policymakers, presented at conferences, and published in peer-reviewed journals to ensure that findings are accessible to the broader scientific and medical communities.

**Trial registration number:** NCT05614349

**Strengths and Limitations Box:**

1. The CanTreatCOVID community-focused design allows enrollment without in-person visits.
2. The adaptive platform trial structure provides flexibility to add promising therapies and remove ineffective ones, which is critical in a rapidly changing pandemic environment
3. CanTreatCOVID gathers real-world data on outpatient COVID-19 care
4. The open-label design avoids logistical challenges associated with placebo controls in large-scale trials, though it may introduce bias related to subjective outcomes
5. The reliance on self-reported adherence to study medications could lead to variability

## INTRODUCTION

While the acute phase of the COVID-19 pandemic has ended, SARS-CoV-2 continues to circulate globally, with over seven million deaths annually attributed to the virus.^1^ Vaccines and natural immunity have now substantially reduced hospitalizations and the need for critical care; however, SARS-CoV-2 is endemic and remains a threat to health.^2^ New variants of SARS-CoV-2 continue to emerge with the risk that vaccine effectiveness and acquired immunity may decline.^3^ Therefore, there will remain an ongoing need for therapeutics that are effective, safe, affordable, and suitable for use in community settings.^4^

While several agents have been evaluated for mild to moderate infections in outpatient settings, current guidelines from Canada, ^5,6^ the United states,^7^ and international bodies^8,9^ primarily have recommended nirmatrelvir/ritonavir (Paxlovid™).^9^ Other therapeutics, such as fluvoxamine or metformin^10^ have moderate (and mixed) supporting evidence,^11^ and inhaled therapeutics have weak evidence to support their use but may be considered.^9,12^ Nirmatrelvir/ritonavir is a promising treatment; however, the key randomized controlled trial (RCT) that assessed its efficacy, the Evaluation of Protease Inhibition for COVID-19 in High-Risk Patients (EPIC-HR) study, was conducted in unvaccinated patients before the emergence of more recent variants of SARS-CoV-2.^13^ Data from the EPIC-SR study showed no significant difference in time to recovery (12 days for nirmatrelvir/ritonavir vs. 13 days for placebo) or hospitalization/death rates (0.8% vs. 1.6%) in vaccinated or unvaccinated non-hospitalized patients with SARS-CoV-2.^14^ A recent systematic review and network meta-analysis of antivirals for non-severe SARS-CoV-2 infection highlighted the need for further studies to evaluate nirmatrelvir/ritonavir and other therapeutics in vaccinated populations and against newer variants of SARS-CoV-2.^15^ In addition, current antiviral therapies for SARS-CoV-2 infection have limitations, including potential adverse effects ranging from mild symptoms to severe reactions, and eligibility restrictions that limit their use in certain populations. In a systematic review we completed, antioxidant treatments hold promise, but further evidence from randomized controlled trials are needed.^16^

Adaptive platform trials (APTs) are an ideal design to assess multiple therapies simultaneously, with the flexibility to introduce new treatments as they emerge.^17^APTs have played a crucial role in identifying effective treatments for hospitalized COVID-19 patients.^18^ RECOVERY identified dexamethasone as significantly reducing mortality in patients requiring oxygen,^19^ and tocilizumab in severe disease.^20^ REMAP-CAP highlighted the efficacy of intravenous steroids in intensive care unit (ICU) patients.^21^ Collaborative efforts across APTs, such as ATTACC, REMAP-CAP, and ACTIV-IV, demonstrated that therapeutic-dose heparin reduces progression to ICU-level care and mortality in non-critically ill hospitalized patients.^22,23^ In the outpatient setting, the PRINCIPLE trial in the UK demonstrated the efficacy of inhaled budesonide.^24^

Given the evolving nature of the virus and the limitations of current therapeutic options, ongoing evaluation of treatments in community settings remains essential to ensure optimal care for patients. The Canadian Adaptive Platform Trial of Treatments for COVID in Community Settings (CanTreatCOVID) is a multi-arm adaptive platform trial that aims to evaluate the clinical and cost-effectiveness of multiple therapeutic interventions compared to usual care for non-hospitalized patients with SARS-COV-2 at high risk of severe complications. The trial uses a flexible design that allows for the assessment of various treatments simultaneously, ensuring that new and promising therapies can be quickly tested and incorporated to improve patient outcomes in community-based settings.

## METHODS AND ANALYSIS

### Trial Design

CanTreatCOVID is an open label, individually randomized, adaptive platform trial conducted across Canada. The trial currently involves six provinces: Ontario, Québec, Alberta, Manitoba, British Columbia, and Newfoundland and Labrador. Each province has a dedicated research hub with a team of research staff, and recruitment occurs through “spokes” in primary care centers and other community settings, as well as other recruitment methods. The primary objective is to evaluate the clinical and cost-effectiveness of multiple therapeutics for non-hospitalized SARS-CoV-2 patients at high risk of severe complications. The trial also aims to address practical challenges related to the implementation of these therapies in diverse healthcare settings. By using an APT design, CanTreatCOVID allows for the continuous evaluation and comparison of treatments, incorporating new interventions as evidence emerges.

### Recruitment strategy

We recruit participants through several channels to ensure broad outreach across Canada. Public communications and community partnerships has played a key role, with recruitment materials available in English and French in each province. The trial website (www.CanTreatCOVID.org) provides clear information and supports public engagement, transparency, and knowledge sharing, based on insights from previous Canadian trials and the PANORAMIC study in the UK^25^. Additionally, a toll-free hotline, available from 8 AM to 6 PM ET, Monday to Friday, provides direct support for potential participants, with trained research assistants available to intake participants and answer questions. We have also developed recruitment materials for public advertising (e.g., transit, print/online media, traditional media) and digital channels (e.g. Google ads, Facebook, Twitter, Instagram).

Primary care practice-based research networks are another source of recruitment. We have engaged primary care practices and physicians, and with their permission, we use existing electronic medical record data to identify patients who meet key eligibility criteria (age and underlying health condition). This has enabled proactive communication with potentially eligible participants should they contract SARS-CoV-2. In outpatient settings, including primary care clinics, urgent care centers, specialty clinics, emergency departments, and pharmacies, we disseminate recruitment materials and encourage referrals, particularly for high-risk populations.

We have leveraged institutional and provincial databases to maximize outreach. Our interdisciplinary team is affiliated with institutions that maintain databases of participants from past studies who have consented to be contacted for future research. This has allowed us to connect directly with individuals who have already expressed a willingness to contribute to scientific studies. Additionally, we collaborate with provincial organizations, such as provincial Ministries of Health, to access databases of individuals who have received the COVID-19 vaccine and consented to be contacted for research purposes.

### Eligibility criteria

Eligible participants include individuals aged 50 years and older, or those aged 18-49 with pre-existing comorbidities that increase their risk of severe illness. Participants must also have a positive SARS-CoV-2 test (PCR or RAT) with proof of a positive test provided via a picture of the result. Patients must be enrolled within 5 days of onset of symptoms associated with SARS-CoV-2 infection^9,26^ because some therapeutics must be taken early in an infection to be effective. Eligible symptoms include at least one of the following: fever or chills, cough, shortness of breath, decreased or loss of taste or smell, runny nose or nasal congestion, headache, fatigue, sore throat, muscle aches or joint pain, or gastrointestinal symptoms.

Exclusion criteria include participants who have been hospitalized or remained in an emergency department for more than 24 hours, prior enrollment in CanTreatCOVID, current participation in another SARS-CoV-2 therapeutic trial, use of or contraindications to a trial therapeutic, or inability of the participant or caregiver to provide informed consent. Patients with contraindications to specific trial drugs are excluded from that particular intervention arm but remain eligible for randomization into other treatment arms. To participate, patients must be eligible for both the usual care arm and at least one intervention arm of the trial.

### Screening

Regardless of how the potential participant learned about the trial, potential participants must speak to a research assistant via telephone call to undergo initial screening. The initial screening process includes reviewing the eligibility criteria and conducting a preliminary assessment of the compatibility of the potential participant’s medications based on a list provided by the participant. The initial screen also serves the potential participant by clarifying questions about trial involvement, and if English is not the first language, research assistants have access to translation services to ensure potential participants can be served without a language barrier. If eligible, the research assistant proceeds to engage the potential participant in the process of obtaining written informed consent. The participant also provides consent to having a team member contact their usual community pharmacy and primary care provider, as needed, to clarify any discrepancies in their health status, including their medications. Once these consents are obtained, the study pharmacist conducts a detailed review of the participant’s medications using a medication list from the patient’s usual pharmacy or provincial repository. If additional information is required to determine eligibility, this is obtained using electronic clinical resources and, if further necessary, the study team contacts the participant’s primary care provider for further clarification. Once the detailed medication and eligibility review has been satisfactorily completed, the study pharmacist makes a recommendation (which may include strategies to mitigate potential drug-interactions) to the provincial principal investigator (PI) who approves and signs off on the participant’s eligibility.

### Randomization and allocation

Following confirmation of eligibility, participants are immediately randomized through an interactive, secure, web-based system maintained by the data management group. Randomization is conducted using fixed, equal allocation ratios corresponding to the number of active treatment arms for which the participant is eligible. If a participant is eligible for two arms (e.g., usual care and one therapeutic), they will be randomized in a 1:1 ratio between these two options. If a participant qualifies for more than two arms, the randomization ratio will adjust accordingly (e.g., 1:1:1 if there are three eligible arms). Initially, participants were randomized in a 1:1 ratio between usual care and the first therapeutic arm (nirmatrelvir/ritonavir). As additional therapeutics are introduced (antioxidant therapy as the second arm), the randomization ratio adjusts accordingly (e.g., 1:1:1). To ensure balance and minimize bias, randomization is stratified by age group (<65 years vs. ≥65 years) and employs permuted blocks of varying sizes.

### Blinding

In this open-label adaptive platform trial, both participants and recruiting clinicians are aware of the assigned intervention. Therefore, no unblinding or code breaking is required. However, trial investigators who are not involved in participant recruitment, as well as the lead statisticians, remain blinded to treatment allocations. Unblinded statisticians, along with independent members of the DSMC, have access to unblinded data. These individuals are responsible for monitoring interim outcomes and will only disclose unblinded information once a decision is reached to stop recruitment for any specific intervention arm.

### Intervention

As the trial progresses, the selection of therapeutics to be evaluated for SARS-CoV-2 treatment in outpatient settings is guided by the Canadian COVID-19 Out-Patient Therapeutics Committee. This committee continuously reviews the latest evidence and provides recommendations to the Steering Committee. The decision to include a new therapy is based on several key criteria:

- Strong scientific and biological rationale, supported by compelling evidence or successful independent studies (e.g., phase I/II trials) demonstrating efficacy for treating SARS-CoV-2 in outpatient settings.
- Sufficient availability of the therapeutic to ensure consistent supply throughout the trial.
- Scalability of the therapeutic, allowing widespread use if found to be effective.
- The addition of a new arm must not interfere with the completion of ongoing research arms.
- The therapeutic must remain clinically relevant by the time the trial arm is completed.

The adaptive platform design and master protocol structure enable the seamless incorporation of new interventions, ensuring flexibility and responsiveness to emerging evidence. Detailed information for each therapeutic, including dosage, known side effects, adverse reactions, duration of treatment, and existing data, is provided to participants along with a patient handout in lay language to ensure clear understanding.

The first intervention arm is nirmatrelvir/ritonavir (Paxlovid™), consisting of 300 mg nirmatrelvir (two 150 mg tablets) with 100 mg ritonavir (one 100 mg tablet), with all three tablets taken together orally twice daily for 5 days. Upon randomization to this arm, participants are urgently provided with a participant pack containing nirmatrelvir/ritonavir, along with dosing instructions and safety information. Participants are advised to complete the full 5-day treatment course, which should be initiated as soon as possible after a diagnosis of SARS-CoV-2 has been made, and ideally within 5 days of symptom onset. Adherence to the study therapeutic is monitored through participant self-reporting.

Participants are eligible for the nirmatrelvir/ritonavir arm if they meet all platform-level inclusion criteria and none of the platform-level exclusion criteria. Additional nirmatrelvir/ritonavir specific exclusion criteria include: a history of clinically significant hypersensitivity to nirmatrelvir /ritonavir or its excipients; known hereditary problems of galactose intolerance, total lactase deficiency or glucose-galactose malabsorption; severe liver impairment (characterized by severe ascites, encephalopathy, jaundice, or prolonged INR (patients with liver disease without any of these features are eligible); being a solid organ transplant recipient on immunosuppressants; having moderate or severe renal disease (defined as CKD stage 3, 4 or 5 or current acute kidney injury or most recent eGFR in the past 6 months <60 ml/min); currently taking nirmatrelvir/ritonavir; clinical requirement to continue taking a drug contraindicated or not recommended for administration with nirmatrelvir/ritonavir; known or suspected pregnancy; breastfeeding; or being of childbearing potential without using highly effective contraception.

The second intervention arm is antioxidant therapy (selenium 300 µg, zinc 40 mg, lycopene 45 mg, and vitamin C 1.5 g) administered as three capsules taken once per day for 10 days. Participants are advised to complete the full 10-day treatment course. Upon randomization to this arm, participants are promptly provided with a participant pack containing the antioxidant therapy, along with dosing instructions and safety information.

Eligibility for the antioxidant therapy arm also requires meeting all platform-level inclusion criteria and none of the platform-level exclusion criteria. Additional exclusion criteria specific to antioxidant therapy include: known or suspected pregnancy, breastfeeding, childbearing potential without highly effective contraception, allergy or intolerance to any of the ingredients (selenium, zinc, lycopene, vitamin C, ascorbyl palmitate, hypromellose, microcrystalline cellulose, or sodium stearyl fumarate), use of warfarin as a preventive measure, advanced chronic kidney disease (CKD stage 3-5, eGFR <60 mL/min), liver disease awaiting transplantation, a history of calcium oxalate kidney stones, head and neck cancer within the past five years, history of non-melanoma skin cancer, current use of supplements with selenium (≥300 µg/day), zinc (≥40 mg/day), lycopene (≥45 mg/day), and vitamin C (≥1500 mg/day), and consumption of omega-3 fatty acids at baseline without willingness to stop during the intervention period.

### Primary and secondary outcomes

Our primary outcome is all-cause hospitalization or death within 28 days from randomization. This is captured through daily diaries (completed from day 1-day 14) as well as during participant follow-up at 21 and 28 days. It will also be cross-checked with administrative data. Based on a robust understanding of SARS-CoV-2 infection, it is likely that severe outcomes would occur within 28 days of symptom onset, ^38^ and this outcome has been used in several key studies of SARS-CoV-2 treatments in community settings.^39–41^ This timeframe also corresponds to when hospitalizations due to adverse drug events from therapy (or drug-interactions) would be expected to occur.

The secondary outcomes include: (1) time to recovery, defined as the first instance that a participant report feeling fully recovered. This will be captured through daily diaries (day 1-day 14) using the questions: “Do you feel recovered today? i.e., symptoms associated with illness are no longer a problem”, which has been used in PRINCIPLE^39^ and PANORAMIC^36^, and the Flu Pro Plus relevant questions about returning to usual health and activities^42^; (2) time to sustained resolution, defined as the number of days from randomization to the first of 3 consecutive days of resolution or alleviation (without subsequent relapse by day 28); (3) time to progression of signs or symptoms, defined as the number of days from randomization to the first of 2 consecutive days of worsening); (4) symptom severity, will be captured through daily diaries using the questions: “How well are you feeling today? Please rate how you are feeling now using a scale of 1 – 4, where 1 is no symptoms, and 4 is very severe symptoms” and by rating symptoms, if present, as “No problem, mild problem, moderate problem, or major problem”; (5) post-acute sequelae of SARS-CoV-2, collected at 90 days and 36 weeks, using the World Health Organization clinical case definition^43,44^ and the validated Symptom Burden Questionnaire for Long COVID^45^; (6) Quality of life, using EQ-5D-5L, collected at baseline, 21 days, 28 days, 90 days, and 36 weeks and analyzed using Canadian reference values; health service use, treatment costs, cost/QALY, analyzed using standard trial-based economic evaluation methods^46^; and (7) early discontinuation and severe adverse events (SAEs).

### Data collection and participants follow up procedures

At baseline, we collect data on sociodemographic including date of birth, sex assigned at birth, gender identity, education level, household income, income source, ethnicity, rurality. We also collect health care numbers, all phone numbers, an emergency contact, an email address, and seek permission to use text messaging, as well as alternative contacts and caregivers (for obtaining follow-up data should the need arise).

All participants receive a call from the trial team 1 day after randomization to confirm the receipt of study materials and medication (if randomized to a study drug) and to address any questions. For participants randomized to nirmatrelvir/ritonavir, an additional safety call is made on day 4 to monitor for any early side effects and to ensure that no new medications have been taken that may be contraindicated with nirmatrelvir/ritonavir. Similarly, participants randomized to the antioxidant therapy arm receive an additional safety call on day 6 to check for potential adverse drug reactions or any contraindicated medications. These safety calls ensure that participants remain safe while adhering to the study protocol.

Participants in all arms will complete an online daily diary each day for 14 days (<u>Supplementary file 1</u>), a validated approach from similar trials.^24,25,27^ Participants self-report symptoms and severity, as well as contact with health services (e.g., hospital admissions, ED visits, visits to specialists and primary care), which will be corroborated with administrative data when the data become available. Moreover, 10% of participants are randomized to complete additional Flu Pro Plus questions in the daily diary, which collect detailed information on severity of symptoms (<u>Supplementary file</u> 2). Our primary study outcome is reported by the patient or their alternative contact and does not rely on administrative data for publication and timely dissemination of results.

Participants are prompted to complete online follow-up surveys using REDCap Cloud© electronically through automated emails sent through the survey software. Research staff call participants with no internet access, as well as those who prefer to complete surveys over the phone. Participants who have not completed their diary for at least two consecutive days before day 7 or again before day 14 will be contacted by staff by email, phone, or text. Staff make 3 attempts over 3 days before considering it missing data.

Research staff contact patients at 21 days and 28 days, focusing on the primary outcome, and at 90 days and 36 weeks from randomization, with a focus on post-acute sequelae of SARS-CoV-2. The EQ-5D-5L^28^ will be administered at baseline, 21 days, 28 days, 90 days, and 36 weeks. Adverse events will be assessed throughout the study period; data on serious adverse events considered by the investigator to be related to the assigned regimen will be collected through the end of study participation.

### Statistical Analysis

#### Primary analysis population

The primary intention-to-treat (ITT) analysis population is defined as all randomised participants according to the group they were randomly allocated to, regardless of deviation from protocol. The ITT Population is typically the primary population for the analysis of efficacy parameters. A subset of efficacy parameters will be evaluated for the per protocol (PP) population, defined as randomized patients who adhered to more than 80% of the assigned therapy. For patients who have an early outcome, 80% applies to the expected number of doses by that time and not the total number. For the usual care group, per protocol will be defined as not receiving an investigational product for the duration corresponding to the PP duration needed for the investigational product. The Safety Population, defined as randomized patients who received at least 1 dose of study drug, is the primary population for the analysis of safety endpoints, compared to the usual care group.

#### Primary and Secondary outcome(s) analysis

For our primary outcome analysis, we use a Bayesian logistic regression model which includes the randomized treatment group, age, and vaccination status as covariates. We will calculate and report model-based estimates of the difference in hospitalization/deaths rates between each treatment arm and the Usual Care, along with 95% Bayesian credible intervals. For the secondary endpoints, logistic regression, ordinal logistic regression, and linear regression will be used, depending on the data types of endpoints. Randomized group, age and vaccination status will be included covariates. Where logistic regression and ordinal logistic regression are used, the odds ratios will be reported for each pairwise treatment arm comparison with the usual care group, along with their respective p-value and 95% confidence interval. Where linear regression is used, the mean and standard deviation will be reported in each treatment arm. The adjusted difference in means, along with the 95% confidence interval for each pairwise treatment arm comparison with the usual care group, will be reported.

The interim analysis plan will be determined by using a series of clinical trial simulations to ensure that satisfactory operating characteristics in terms of controlling the type I error rate in cases where all treatment arms are the same, while attempting to maximize the statistical power in cases where one or more treatment arms yield meaningful treatment benefits. Before any unblinded analyses are performed by an independent statistician, the simulations will be performed by the Methods and Statistical Analysis Committee who will be blinded to the knowledge of group assignments.

The interim analysis will determine a) the superiority of the treatment arm in comparison with the Standard of Care (SOC); b) switching the SOC arm to another treatment arm with significant superiority; c) futility and suspension of a treatment arm; and d) adding additional treatment arms.

Interim futility determines whether a treatment arm should be suspended, i.e., halted from receiving new patients for the next cohort. We compute the model-based hospitalization/death rate for each treatment arm, denoted as p_j_, with the stratification covariates being set to the respective populational averages. Using Markov chain Monte Carlo (MCMC) sampling, we compute the Bayesian posterior probability that hospitalization/death rate of treatment arm j is smaller than that of the SOC by at least a margin of 1%, Pr(p_SOC_ - p_j_ > 0.01), and if it is smaller than a precalibrated stopping boundary, e.g., 0.001, the interim futility rule for hospitalization/death endpoint is satisfied. If interim futility rules are satisfied for a treatment, then it will be suspended over the next cohort. If the two interim futility rules are no longer satisfied in the next interim analysis in the light of new data in other treatment arms, the suspension might be canceled.

Subgroup analyses will be performed on several populations, including age, sex, gender, body mass index (BMI), income, race and ethnicity, rurality, medical conditions (multimorbidity and immunocompromised), number of vaccine doses received (< 2 vs ≥2), and place of recruitment (emergency departments vs other settings). We will also examine the moderating effects of concomitant medications, such as corticosteroids. For each subgroup, we will calculate model-based estimates of the difference in rates of emergency visits, death, and hospitalization between the treatment arms and the usual care, accompanied by 95% Bayesian credible intervals computed using the primary analysis model.

### Sample size

Given the open, ‘perpetual’ trial structure, the trial does not have a finite ending based on sample size. We have determined our estimated sample size per arm based on Bayesian logistic regression to estimate the adjusted odds ratio for hospitalization or death at 28 days for a pairwise comparison of treatment arm (initially nirmatrelvir/ritonavir) vs. comparator (initially usual care). Assuming a conservative 5% comparator event rate of hospitalization or death^29,30^, we deemed that a reduction from 5% to 3.35% would be clinically meaningful, based on the current published evidence.^13,24,31^ We estimate that approximately 2,413 participants per arm will be required to provide 90% power for detecting a 25% reduction in the relative risk of hospitalization/death.

### Ethical considerations and governance

The CanTreatCOVID master protocol and sub-protocols have been approved by Health Canada and local research ethics boards in the participating provinces across Canada. All participants provide informed consent, which can be completed online, via email, or verbally via telephone. An independent Data Monitoring and Safety Committee (DMSC) oversees safety data, review interim analyses provided by the Statistical Analysis Committee (SAC), and communicate recommendations to the Trial Steering Committee (TSC). The TSC provides guidance and oversight to the Trial Management Group (TMG). The results of the study will be disseminated to policymakers, presented at conferences, and published in peer-reviewed journals to ensure that findings are accessible to the broader scientific and medical communities.

## DISCUSSION

CanTreatCOVID is an open-label, individually randomized, adaptive platform trial designed to evaluate therapeutics for SARS-CoV-2 infection in high-risk outpatient populations. Eligible participants include those aged 50 years or older, or those aged 18-49 with at least one chronic medical condition or immunosuppression. Patients must be enrolled within 5 days of symptom onset. The trial evaluates therapeutics against usual care and may compare treatments directly as new interventions are added. The Canadian COVID-19 Out-Patient Therapeutics Committee oversees the selection of investigational therapies based on emerging evidence, ensuring that relevant and promising treatments are assessed in real-time.

CanTreatCOVID’s adaptive design allows modifications based on interim analyses of primary outcome data, enabling the removal of interventions deemed futile or successful. This structure also facilitates the integration of findings from international trials and the addition of new treatment arms as they become available. This flexible approach ensures the trial remains responsive to the evolving nature of the COVID-19 pandemic and the therapeutic landscape, while contributing to the broader knowledge base on outpatient management of SARS-CoV-2.

### Comparison with other studies

Several other adaptive platform trials (APTs) share similar objectives and methodologies. The multi-center, adaptive, randomized platform trial to evaluate the effect of repurposed medicines in outpatients with early COVID-19 and high-risk for complications (TOGETHER) trial ^32^ is multi-center APT assesses repurposed medicines for treating high-risk COVID-19 outpatients in early stages of infection. The study was conducted in Brazil and recruited over 10,000 participants. The trial has concluded investigations on six medications including hydroxychloroquine, lopinavir / ritonavir, ivermectin, fluvoxamine maleate, metformin, and doxazosin; and currently have four treatments (Peginterferon Lambda, fluvoxamine, fluvoxamine plus molnupiravir and fluvoxamine plus inhaled corticosteroid) under investigation. The trial ability to test a range of inexpensive, repurposed drugs has offered valuable data on treatments that are scalable, especially in low-and middle-income countries. While TOGETHER shares CanTreatCOVID’s focus on early treatment in outpatients, it differs in its geographic scope and focus on repurposed drugs.

The Platform Adaptive trial of NOvel antiviRals for eArly treatment of COVID-19 in the Community (PANORAMIC)^31^ conducted in the UK, is one of the largest APTs for outpatient COVID-19 therapeutics. It is an open-label APT that assessed the safety and efficacy of antiviral treatments versus usual care in patients at increased risk of COVID-19 related morbidity and mortality. PANORAMIC has enrolled over 29,000 participants across 65 sites. The trial evaluated both molnupiravir^25^ and nirmatrelvir/ritonavir. Recruitment for both treatments has now concluded, with the trial playing a significant role in confirming the real-world effectiveness of these antiviral agents in highly vaccinated populations. PANORAMIC shares CanTreatCOVID’s community-based approach, but it is primary focused on antiviral treatments, while CanTreatCOVID aims to evaluate a broader range of therapies.

The Accelerating COVID-19 Treatment Interventions and Vaccines (ACTIV-2 and ACTIV-6) were launched by the U.S. National Institutes of Health to assess outpatient treatments for mild-to-moderate COVID-19 cases in individuals at risk of severe disease progression.^33^ ACTIV-2, a Phase II/III trial, evaluates monoclonal antibodies, oral antivirals, and inhaled interferons and has enrolled 4,044 participants across 172 sites worldwide, including locations in Argentina, Brazil, and the United States.^33,34^ ACTIV-6, a Phase III platform trial, examines the effectiveness of widely available prescription and over-the-counter medications in managing COVID-19 symptoms in outpatients.^33^ With a focus on repurposed medications, such as ivermectin and fluvoxamine, ACTIV-6 has enrolled over 10,000 participants across 110 sites within the United States.^33^ Similar to TOGETHER, ACTIV-6 aims to provide data on accessible, affordable treatments, but focuses on medications readily available to the public. Both ACTIV-2 and ACTIV-6 provide valuable insights into outpatient management of SARS-CoV-2, though they differ from CanTreatCOVID by targeting specific therapeutic classes rather than a broad spectrum of treatments.

### Strengths and limitations

One of CanTreatCOVID’s major strengths is its community-focused approach, which prioritizes early intervention for patients in outpatient settings. By allowing enrollment without the need for in-person visits, the trial maximizes accessibility and reduces barriers to participation. This approach captures a broader, more diverse patient population, and also ensures that therapeutics are tested during the early, milder stages of COVID-19, where timely intervention may yield the greatest benefits. This emphasis on community care reflects the growing recognition that managing SARS-CoV-2 in outpatient settings is critical to reducing the burden on healthcare systems and preventing severe outcomes. Another key strength is the trial’s adaptive design, which allows for real-time flexibility. As new data become available, treatments that show promise can be added, while ineffective ones can be removed. This adaptability is essential in a rapidly evolving pandemic, where new variants and therapies continue to emerge.

A key limitation of the trial is its open-label design. Without blinding, both participants and study clinicians are aware of the intervention being administered, which introduces the potential for bias. However, the decision to forgo placebo control was made deliberately, as usual care in the community does not typically include placebo administration. Additionally, creating placebo equivalents for multiple investigational drugs in a large-scale platform trial would be logistically challenging and may delay the rapid evaluation of potentially life-saving treatments. While the open-label design could impact subjective outcomes, it remains the most feasible and practical option given the trial’s scale and the urgency of the pandemic. Furthermore, previous open-label trials have shown no difference in subjective outcomes, specifically, time to recovery between different intervention arms and usual care^35^, indicating that the absence of a placebo did not impact participants’ perception of being fully recovered.

Another potential limitation is the reliance on self-reported data for adherence to the study medications, which may introduce variability in how accurately participants follow the therapeutic regimen. To address this, the trial uses online questionnaires and regular check-ins with study staff to support accurate reporting and help participants stick to the regimen.

CanTreatCOVID represents a robust, adaptive platform trial designed to address the urgent need for effective therapeutics in the outpatient management of SARS-CoV-2. By focusing on high-risk populations and leveraging real-world healthcare settings, the trial offers the potential to identify treatments that can significantly improve outcomes in the community. The adaptive design enables the trial to evolve in response to emerging evidence. While challenges such as the open-label design and self-reported adherence exist, these are outweighed by the trial’s strengths in accessibility, scalability, and responsiveness to the changing pandemic landscape.

## Data Availability

All data produced in the present study are available upon reasonable request to the authors.

## Contributors

ADP, BH, PD, MG, PJ, TL, EM, SM, PS conceived of the work. BH and ADP drafted the work. All authors contributed to revising the manuscript for important intellectual content, gave final approval of the version to be published and agreed to be accountable for all aspects of the work.

## Acknowledgements

The authors thank our incredible patient and community partners: Brenda Andreas, Cris Carter, Jane Cooney, Gabriela Covaci, Letlotlo Gariba, Jennifer Hulme, Veronika Kiryanova, Kathy Kobow, Mike Lapenna, Mary Liu, Chris Maddison, Dorothy Nelson, Moon Ja Park, Lyric Paul, Donna Rubenstein, Dorothy Senior, Allard Schipper, Kimberly Strain, Margo Twohig, Mike Warren, John Zhan, and Alexander Zsager.

---

**Table 1-.**
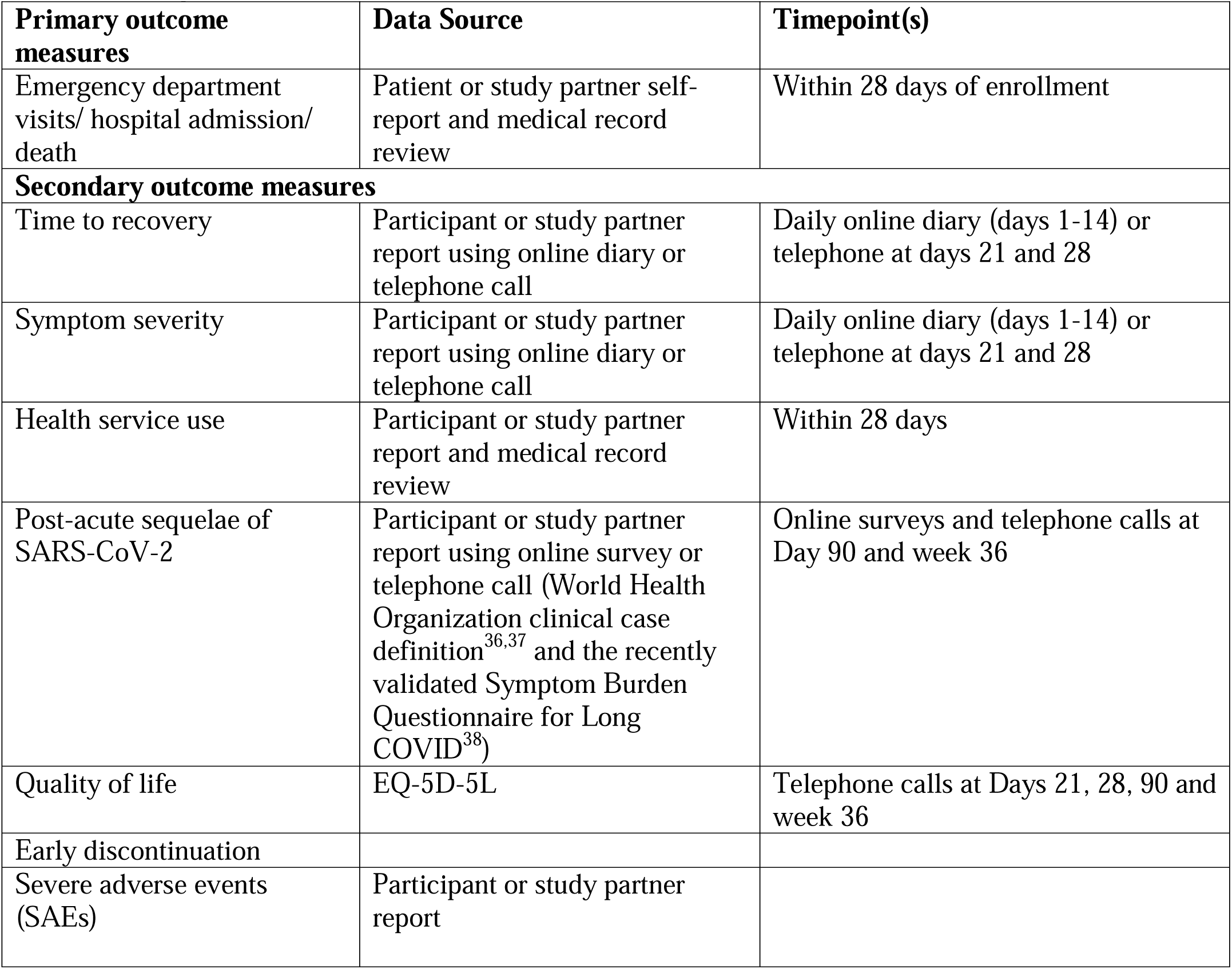
Study outcomes in CanTreatCOVID Trial

| **Table 1- Study outcomes in CanTreatCOVID Trial** |  |  |
| --- | --- | --- |
| **Primary outcome measures** | **Data Source** | **Timepoint(s)** |
| Emergency department visits/ hospital admission/ death | Patient or study partner self-report and medical record review | Within 28 days of enrollment |
| **Secondary outcome measures** |  |  |
| Time to recovery | Participant or study partner report using online diary or telephone call | Daily online diary (days 1-14) or telephone at days 21 and 28 |
| Symptom severity | Participant or study partner report using online diary or telephone call | Daily online diary (days 1-14) or telephone at days 21 and 28 |
| Health service use | Participant or study partner report and medical record review | Within 28 days |
| Post-acute sequelae of SARS-CoV-2 | Participant or study partner report using online survey or telephone call (World Health Organization clinical case definition<sup>36,37</sup> and the recently validated Symptom Burden Questionnaire for Long COVID<sup>38</sup>) | Online surveys and telephone calls at Day 90 and week 36 |
| Quality of life | EQ-5D-5L | Telephone calls at Days 21, 28, 90 and week 36 |
| Early discontinuation |  |  |
| Severe adverse events (SAEs) | Participant or study partner report |  |

## Supplementary File 1: Participant daily dairy

**Date**

Today’s date: _ _/ _ _ _ / (DD/MMM/YYYY)

**Symptoms**

1. Do you feel you have recovered today? (i.e., Symptoms associated with the illness are no longer a problem).

___ YES

___ NO

2. How are you feeling today? 0 no symptoms/ 1 Mild / 2 Moderate/ 3 Severe/ 4 Very severe

3. Please rate interference in daily activities due to illness: 1 Not at all/ 2 A little bit/ 3 Somewhat/ 4 Quite a bit/ 5 Very much

4. How is your general health? 1 Poor/ 2 Fair/ 3 Good / 4 Very good/ 5 Excellent

5. Have you returned to your usual health today?

____ YES

____ No

6. Have you returned to your usual activities today?

____ YES

____ No

6a) For those taking study medication, have you taken the prescribed dose?

____ YES

____ No

6b) If no, why not? _________________________

If you feel recovered and returned to your usual activities, you do not need to answer any further questions today. Thank you for your time.

______________________________

As you do not feel recovered, please rate the following symptoms:

7. Fever: No problem / Mild problem / Moderate problem / Major problem

8. Cough: No problem / Mild problem / Moderate problem / Major problem

9. Shortness of breath: No problem / Mild problem / Moderate problem / Major problem

10. Loss of taste/ smell: No problem / Mild problem / Moderate problem / Major problem

11. Muscle ache: No problem / Mild problem / Moderate problem / Major problem

12. Nausea / vomiting: No problem / Mild problem / Moderate problem / Major problem

13. Fatigue: No problem / Mild problem / Moderate problem / Major problem

14. Difficult concentrating: No problem / Mild problem / Moderate problem / Major problem

15. Anxious mood: No problem / Mild problem / Moderate problem / Major problem

16. Please describe any other symptoms with your current illness:

_______________________________________________________________________________

_______________________________________________________________________________

_______________________________________________________________________________

17. Please tell us whether or not you have taken any of these following today. Please answer Yes or No.

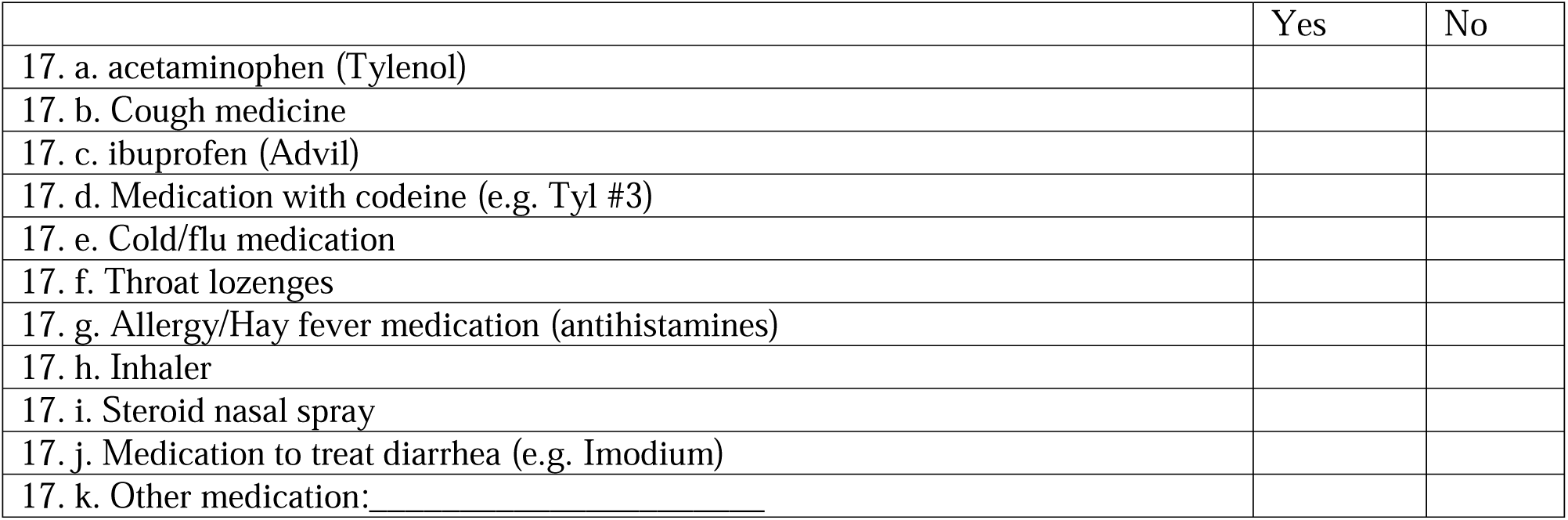

**Healthcare services**

18. Have you contacted or visited the following healthcare services in the last 24 hours? Please answer Yes or No.

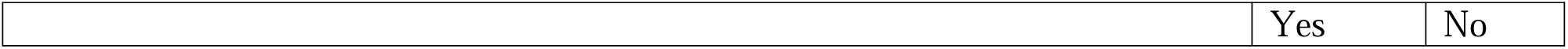
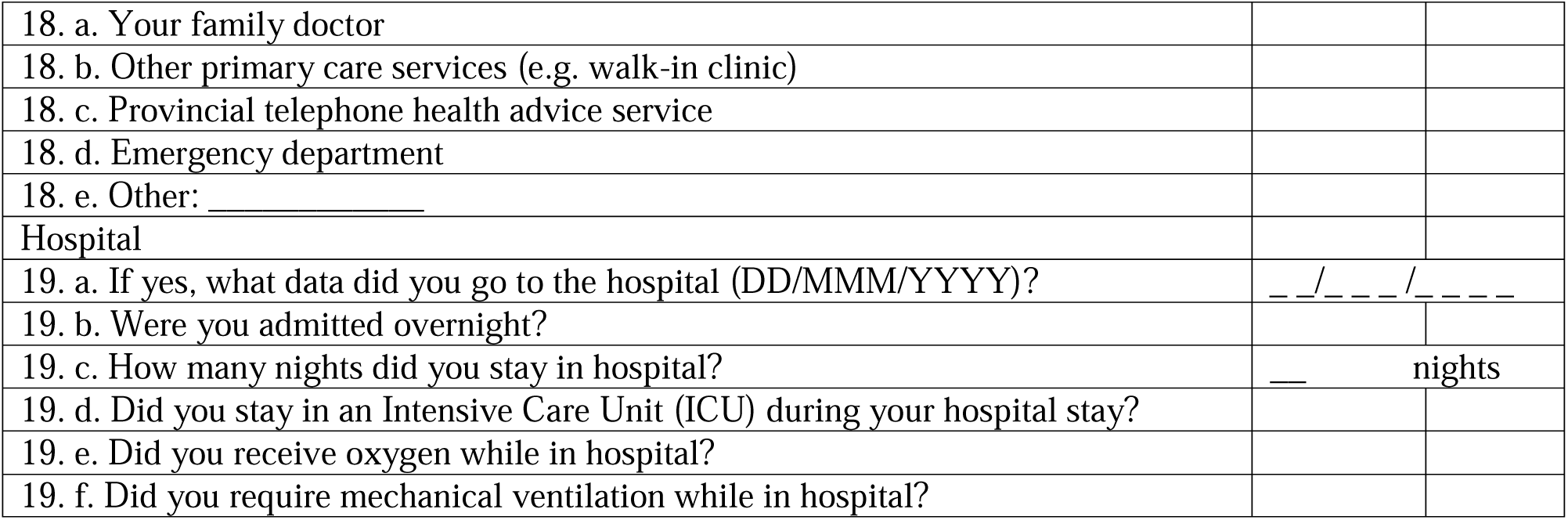

### Supplementary File 2: Participant daily dairy (Flu Pro Plus)

**Date**

Today’s date: _ _/ _ _ _ / (DD/MMM/YYYY)

**Symptoms**

7. Do you feel you have recovered today? (i.e., Symptoms associated with the illness are no longer a problem).

____ YES

____ No

8. How are you feeling today? 0 no symptoms/ 1 Mild / 2 Moderate/ 3 Severe/ 4 Very severe

9. Please rate interference in daily activities due to illness: 1 Not at all/ 2 A little bit/ 3 Somewhat/ 4 Quite a bit/ 5 Very much

10. How is your general health? 1 Poor/ 2 Fair/ 3 Good / 4 Very good/ 5 Excellent

11. Have you returned to your usual health today?

____ YES

____ No

12. Have you returned to your usual activities today?

____ YES

____ No

6a) For those taking study medication, have you taken the prescribed dose?

____ YES

____ No

6b) If no, why not? ____________________________

If you feel recovered and returned to your usual activities, you do not need to answer any further questions today. Thank you for your time.

________________________________________________________

As you do not feel recovered, please rate the following symptoms:

7. Fever: No problem / Mild problem / Moderate problem / Major problem

8. Cough: No problem / Mild problem / Moderate problem / Major problem

9. Shortness of breath: No problem / Mild problem / Moderate problem / Major problem

10. Loss of taste/ smell: No problem / Mild problem / Moderate problem / Major problem

11. Muscle ache: No problem / Mild problem / Moderate problem / Major problem

12. Nausea / vomiting: No problem / Mild problem / Moderate problem / Major problem

13. Fatigue: No problem / Mild problem / Moderate problem / Major problem

14. Difficult concentrating: No problem / Mild problem / Moderate problem / Major problem

15. Anxious mood: No problem / Mild problem / Moderate problem / Major problem

***Flu-Pro Plus***

**Nose**

16. Runny or dripping Not at all/ A little bit/ Somewhat/ Quite a bit/ Very much

17. Congestion or stuffy Not at all/ A little bit/ Somewhat/ Quite a bit/ Very much

18. Sneezing Not at all/ A little bit/ Somewhat/ Quite a bit/ Very much

19. Sinus pressure Not at all/ A little bit/ Somewhat/ Quite a bit/ Very much

**Throat**

20. Sore throat Not at all/ A little bit/ Somewhat/ Quite a bit/ Very much

21. Scratchy or itchy throat Not at all/ A little bit/ Somewhat/ Quite a bit/ Very much

22. Difficulty swallowing Not at all/ A little bit/ Somewhat/ Quite a bit/ Very much

**Eyes**

23. Teary or watery eyes Not at all/ A little bit/ Somewhat/ Quite a bit/ Very much

24. Sore or painful eyes Not at all/ A little bit/ Somewhat/ Quite a bit/ Very much

25. Eyes sensitive to light Not at all/ A little bit/ Somewhat/ Quite a bit/ Very much

**Chest/Respiratory**

26. Trouble breathing Not at all/ A little bit/ Somewhat/ Quite a bit/ Very much

27. Chest congestion Not at all/ A little bit/ Somewhat/ Quite a bit/ Very much

28. Chest tightness Not at all/ A little bit/ Somewhat/ Quite a bit/ Very much

29. Dry or hacking cough Not at all/ A little bit/ Somewhat/ Quite a bit/ Very much

30. Wet or loose cough Not at all/ A little bit/ Somewhat/ Quite a bit/ Very/much

42. Body aches or pains Not at all/ A little bit/ Somewhat/ Quite a bit/ Very much

43. Weak or tired Not at all/ A little bit/ Somewhat/ Quite a bit/ Very much

44. Chills of shivering much

45. Felt cold much

46. Felt hot much

47. Sweating much

**Sense**

48. Lack of taste much

49. Lack of smell much

50. Please describe any other symptoms with your current illness:

_______________________________________________________________________________

_______________________________________________________________________________

_______________________________________________________________________________

51. Please tell us whether or not you have taken any of these following today. Please answer Yes or No.

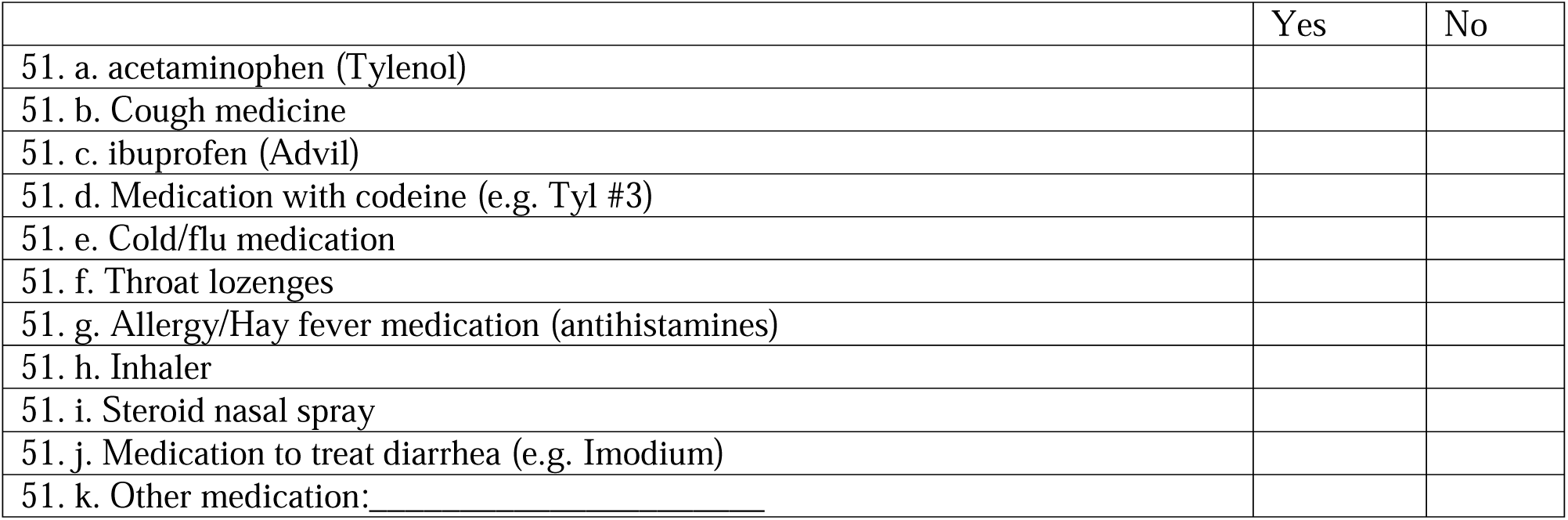

**Healthcare services**

52. Have you contacted or visited the following healthcare services in the last 24 hours? Please answer Yes or No.

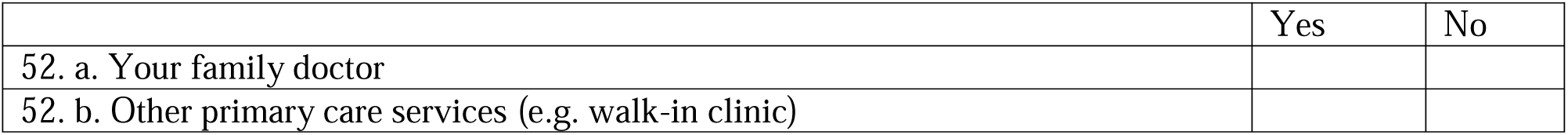
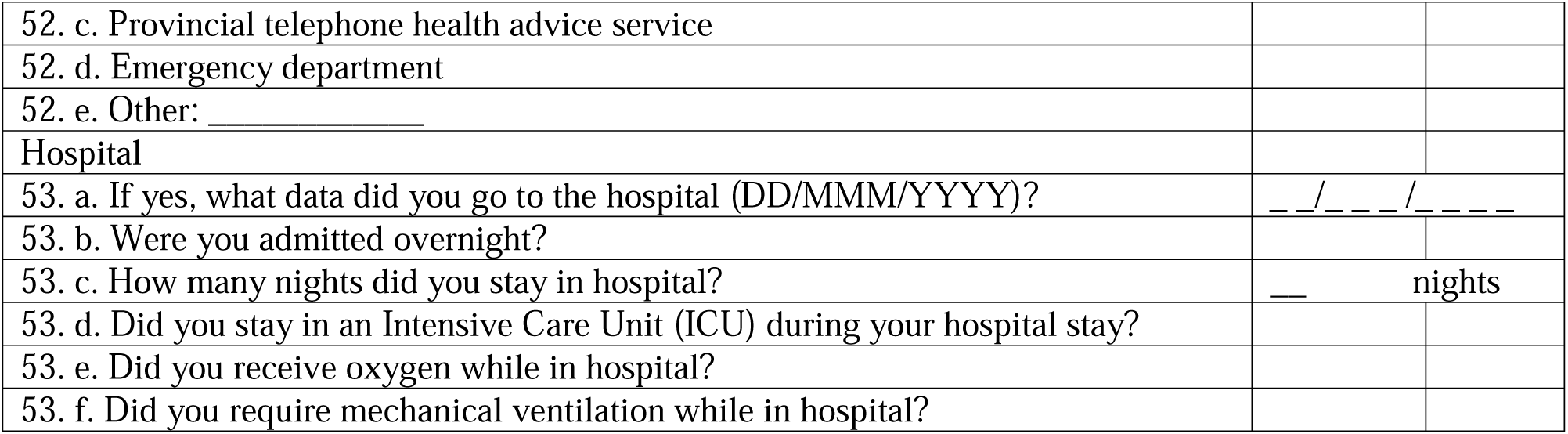

