## Appendix 1 for "Canadian Adaptive Platform Trial of Treatments for COVID in Community Settings (CanTreatCOVID): protocol for a randomized controlled adaptive platform trial of treatments for acute SARS-CoV-2 infection in community settings"

**Date**

Today’s date: _ _/ _ _ _ / _ _ _ _ (DD/MMM/YYYY)

**Symptoms**

1. Do you feel you have recovered today? (i.e., Symptoms associated with the illness are no longer a problem).

____ YES

____ NO

2. How are you feeling today? 0 no symptoms/ 1 Mild / 2 Moderate/ 3 Severe/ 4 Very severe
3. Please rate interference in daily activities due to illness: 1 Not at all/ 2 A little bit/ 3 Somewhat/ 4 Quite a bit/ 5 Very much
4. How is your general health? 1 Poor/ 2 Fair/ 3 Good / 4 Very good/ 5 Excellent
5. Have you returned to your usual health today?

____ YES

____ No

6. Have you returned to your usual activities today?

____ YES

____ NO

6a) For those taking study medication, have you taken the prescribed dose?

____ YES

____ NO

6b) If no, why not? _________________________________________

If you feel recovered and returned to your usual activities, you do not need to answer any further questions today. Thank you for your time.

As you do not feel recovered, please rate the following symptoms:

7. Fever:	No problem / Mild problem / Moderate problem / Major problem

8. Cough:	No problem / Mild problem / Moderate problem / Major problem

9. Shortness of breath:	No problem / Mild problem / Moderate problem / Major problem

10. Loss of taste/ smell: No problem / Mild problem / Moderate problem / Major problem

11. Muscle ache:	No problem / Mild problem / Moderate problem / Major problem

12. Nausea / vomiting:	No problem / Mild problem / Moderate problem / Major problem

13. Fatigue:	No problem / Mild problem / Moderate problem / Major problem

14. Difficult concentrating:	No problem / Mild problem / Moderate problem / Major problem

15. Anxious mood:	No problem / Mild problem / Moderate problem / Major problem

16. Please describe any other symptoms with your current illness:

17. Please tell us whether or not you have taken any of these following today. Please answer Yes or No.

|  | Yes | No |
| --- | --- | --- |
| 17. a. acetaminophen (Tylenol) |  |  |
| 17. b. Cough medicine |  |  |
| 17. c. ibuprofen (Advil) |  |  |
| 17. d. Medication with codeine (e.g. Tyl #3) |  |  |
| 17. e. Cold/flu medication |  |  |
| 17. f. Throat lozenges |  |  |
| 17. g. Allergy/Hay fever medication (antihistamines) |  |  |
| 17. h. Inhaler |  |  |
| 17. i. Steroid nasal spray |  |  |
| 17. j. Medication to treat diarrhea (e.g. Imodium) |  |  |
| 17. k. Other medication:______________________ |  |  |

**Healthcare services**

18. Have you contacted or visited the following healthcare services in the last 24 hours? Please answer Yes or No.

|  | Yes | No |
| --- | --- | --- |
| 18. a. Your family doctor |  |  |
| 18. b. Other primary care services (e.g. walk-in clinic) |  |  |
| 18. c. Provincial telephone health advice service |  |  |
| 18. d. Emergency department |  |  |
| 18. e. Other: ____________ |  |  |
| Hospital |  |  |
| 19. a. If yes, what data did you go to the hospital (DD/MMM/YYYY)? | _ _/_ _ _ /_ _ _ _ | |
| 19. b. Were you admitted overnight? |  |  |
| 19. c. How many nights did you stay in hospital? | __ nights | |
| 19. d. Did you stay in an Intensive Care Unit (ICU) during your hospital stay? |  |  |
| 19. e. Did you receive oxygen while in hospital? |  |  |
| 19. f. Did you require mechanical ventilation while in hospital? |  |  |
