## Appendix 2 for "Canadian Adaptive Platform Trial of Treatments for COVID in Community Settings (CanTreatCOVID): protocol for a randomized controlled adaptive platform trial of treatments for acute SARS-CoV-2 infection in community settings"

**Chest/Respiratory**

26. Trouble breathing	Not at all/	A little bit/	Somewhat/	Quite a bit/ Very much
27. Chest congestion	Not at all/	A little bit/	Somewhat/	Quite a bit/ Very much
28. Chest tightness	Not at all/	A little bit/	Somewhat/	Quite a bit/ Very much
29. Dry or hacking cough	Not at all/	A little bit/	Somewhat/	Quite a bit/ Very much
30. Wet or loose cough	Not at all/	A little bit/	Somewhat/	Quite a bit/ Very much
31. Sputum (coughing up sputum or phlegm) Not at all/	A little bit/	Somewhat/	Quite a bit/ Very much
32. Wheezing	Not at all/	A little bit/	Somewhat/	Quite a bit/ Very much

**Gastrointestinal**

33. Felt nauseous	Not at all/	A little bit/	Somewhat/	Quite a bit/ Very much
34. Stomach ache	Not at all/	A little bit/	Somewhat/	Quite a bit/ Very much
35. Vomit	Not at all/	A little bit/	Somewhat/	Quite a bit/ Very much
36. Diarrhea	Not at all/	A little bit/	Somewhat/	Quite a bit/ Very much

**Body/Systemic**

37. Felt Dizzy	Not at all/	A little bit/	Somewhat/	Quite a bit/ Very much
38. Head congestion	Not at all/	A little bit/	Somewhat/	Quite a bit/ Very much
39. Headache	Not at all/	A little bit/	Somewhat/	Quite a bit/ Very much
40. Lack of appetite	Not at all/	A little bit/	Somewhat/	Quite a bit/ Very much
41. Sleeping more than usual	Not at all/	A little bit/	Somewhat/	Quite a bit/ Very much
42. Body aches or pains	Not at all/	A little bit/	Somewhat/	Quite a bit/ Very much
43. Weak or tired	Not at all/	A little bit/	Somewhat/	Quite a bit/ Very much
44. Chills of shivering	Not at all/	A little bit/	Somewhat/	Quite a bit/ Very much
45. Felt cold	Not at all/	A little bit/	Somewhat/	Quite a bit/ Very much
46. Felt hot	Not at all/	A little bit/	Somewhat/	Quite a bit/ Very much
47. Sweating	Not at all/	A little bit/	Somewhat/	Quite a bit/ Very much

**Sense**

48. Lack of taste	Not at all/	A little bit/	Somewhat/	Quite a bit/ Very much
49. Lack of smell	Not at all/	A little bit/	Somewhat/	Quite a bit/ Very much

50. Please describe any other symptoms with your current illness:

51. Please tell us whether or not you have taken any of these following today. Please answer Yes or No.

|  | Yes | No |
| --- | --- | --- |
| 51. a. acetaminophen (Tylenol) |  |  |
| 51. b. Cough medicine |  |  |
| 51. c. ibuprofen (Advil) |  |  |
| 51. d. Medication with codeine (e.g. Tyl #3) |  |  |
| 51. e. Cold/flu medication |  |  |
| 51. f. Throat lozenges |  |  |
| 51. g. Allergy/Hay fever medication (antihistamines) |  |  |
| 51. h. Inhaler |  |  |
| 51. i. Steroid nasal spray |  |  |
| 51. j. Medication to treat diarrhea (e.g. Imodium) |  |  |
| 51. k. Other medication:______________________ |  |  |

**Healthcare services**

52. Have you contacted or visited the following healthcare services in the last 24 hours? Please answer Yes or No.

|  | Yes | No |
| --- | --- | --- |
| 52. a. Your family doctor |  |  |
| 52. b. Other primary care services (e.g. walk-in clinic) |  |  |
| 52. c. Provincial telephone health advice service |  |  |
| 52. d. Emergency department |  |  |
| 52. e. Other: ____________ |  |  |
| Hospital |  |  |
| 53. a. If yes, what data did you go to the hospital (DD/MMM/YYYY)? | _ _/_ _ _ /_ _ _ _ | |
| 53. b. Were you admitted overnight? |  |  |
| 53. c. How many nights did you stay in hospital? | __ nights | |
| 53. d. Did you stay in an Intensive Care Unit (ICU) during your hospital stay? |  |  |
| 53. e. Did you receive oxygen while in hospital? |  |  |
| 53. f. Did you require mechanical ventilation while in hospital? |  |  |
